# Development and Validation of a Web-based Prediction Model for Acute Kidney Injury after surgery

**DOI:** 10.1101/2020.07.03.20145094

**Authors:** Sang H. Woo, Jillian Zavodnick, Lily Ackermann, Omar Maarouf, Jingjing Zhang, Scott W. Cowan

## Abstract

**Background and objectives:** Acute kidney injury after surgery is associated with high mortality and morbidity. The purpose of this study is to develop and validate a risk prediction tool for the occurrence of postoperative acute kidney injury requiring renal replacement therapy.

**Design, setting, participants, measurements:** This retrospective cohort study had 2,299,502 surgical patients over 2015-2017 from the American College of Surgeons National Surgical Quality Improvement Program Database (ACS-NSQIP). Eleven predictors were selected for the predictive model: age, history of congestive heart failure, diabetes, ascites, emergency surgery, preoperative serum creatinine, hematocrit, sodium, preoperative sepsis, preoperative acute renal failure and surgery type. The predictive model was trained using 2015-2016 data (n=1,487,724) and further tested using 2017 data (n=811,778). A risk model was developed using multivariate logistic regression and machine learning methods.

**Main outcomes:** The primary outcome was postoperative 30-day acute kidney injury requiring renal replacement therapy(AKI-D)

**Results:** The unadjusted 30-day postoperative mortality rate associated with AKI-D was 37.5%. The renal risk prediction model had high AUC (area under the receiver operating characteristic curve, training cohort: 0.89, test cohort: 0.90) for postoperative AKI-D.

**Conclusions:** This model provides a clinically useful bedside predictive tool for postoperative acute kidney injury requiring dialysis.

## INTRODUCTION

Postoperative renal failure is a serious medical complication of surgery, with a rising incidence and significantly associated morbidity and mortality. Postoperative acute kidney injury (AKI) increases five-year mortality regardless of level of injury and extent of kidney recovery. ^1^ For patients who develop acute kidney injury requiring dialysis (AKI-D) after elective surgery, ninety-day mortality rate was reported to be 42%; among patients undergoing cardiac surgery, dialysis conferred a thirty-day mortality rate of 63.7% as compared to 4.3% without AKI. ^2 3^ Of patients who survived the episode of AKI-D, 27.2% required chronic dialysis, with its attendant morbidity and cost.^3^ Hospital costs are significantly higher for patients who develop any postoperative AKI. ^4^

Preoperative assessment of surgical risk is valuable for determining appropriateness of surgery, facilitating multidisciplinary surgical planning, and engaging patients in shared decision-making. ^5^ Tools for cardiac and pulmonary risk stratification have been widely adopted, but the same is not true for renal risk assessment. ^6 7 8 9^ Existing postoperative AKI models are based on small datasets or are specific to patients undergoing cardiac surgery. ^10 11 12,13 14^ The Universal American College of Surgeons (ACS) National Surgical Quality Improvement Program (NSQIP) Surgical Risk Calculator includes postoperative renal failure as one of its 8 outcomes. ^15^ The need to input 21 clinical factors into the calculator limits the adoption of the NSQIP calculator, especially for clinicians who are mainly interested in renal outcome. ^15^

Establishing a renal risk prediction model for surgical patients including the risk for renal replacement therapy is a clinical unmet need. The objective of this study is to develop and validate an AKI-D risk calculator using data from the NSQIP database.

## METHODS

### Study participants

Data were obtained from the ACS NSQIP database, which collects clinical data from more than 500 United States hospitals including patient demographics, medical conditions, laboratory data, and 30-day postoperative outcomes collected by trained surgical clinical reviewers. ^16 17^ Patients undergoing surgery at participating hospitals from 2015 to 2017 were included, with a total population of 2,299,502. The dataset from 2015 to 2016 was used to develop the model (n= 1,487,724), while the dataset from 2017 was held for validation. The study was approved by the Thomas Jefferson University Institutional Review Board and informed consents were waived.

### Outcome

The outcome of interest was AKI-D within thirty days of surgery, a time frame chosen based on the ACS-NSQIP definition. ACS-NSQIP defines AKI-D as “worsening of renal dysfunction postoperatively requiring hemodialysis, peritoneal dialysis, hemofiltration, hemodiafiltration, or ultrafiltration in a patient who did not require dialysis preoperatively.” ^18^ Instructions from the organization state that if a patient refuses a recommendation for dialysis, they should still be coded as developing AKI-D as dialysis was deemed necessary. ^18^

### Data Analysis

We analyzed previously-described risk factors for AKI including demographic factors, medical history, current medical condition (e.g. presence of sepsis, American Society of Anesthesiologists (ASA) class), and preoperative functional status. Multiple preoperative laboratory tests were analyzed including serum sodium, creatinine, platelets, and hematocrit. Participant risk factors were compared using Chi-square (χ^2^) for categorical variables and Wilcoxon for continuous variables. Characteristics of patients who developed AKI-D were compared by adjusted odds ratio. Multivariate logistic regression was performed with risk factors to determine the significance of their associations with the outcome.

Separate categories were made for missing values of laboratory concentrations of serum hematocrit. Backwards elimination and clinical importance were used to select predictor variables of the model. The model was trained on 2015-2016 cohort data and tested on 2017 cohort.

GridSearchCV was used to perform 5 fold cross validation to select the best model parameters. Data set from year 2017 (n=811,778) was used to further test and validate the predictive model. Area under the receiver operating characteristic curve (AUC) was used for model evaluation.

Statistical analysis was performed with Python (version 3.65), Statsmodels (version 0.8.0), and R programming (RStudio version 1.1.463). Scikit-learn was used for machine learning modeling.

## RESULTS

### Study population

The study included 2,299,502 patients from 2015-2017. 56.4 % were women. **Table 1** shows the characteristics of study patients from the years 2015-2016. There were 0.3% of patients who developed AKI-D. Patients who required dialysis were older (66.2 years versus 58.5, p<0.001), had higher preoperative serum creatinine level (1.96 mg/dl vs. 0.92 mg/dl, p<0.001), and were more likely to have congestive heart failure (8.83 % vs. 0.93%, p<0.001), diabetes (34.2% vs 17.19%, p<0.001), or other comorbidities. They were more likely to have undergone emergency surgery (37.8% versus 10.4%, p<0.001). **eFigure 1** in the **Supplement** shows a flow chart of study participants.

**Table 1.** Patient characteristics (2015-2016)

|  | No AKI with Dialysis<br>(n=1,483,169) | AKI with Dialysis<br>(n=4,555) | p-value |
| --- | --- | --- | --- |
| Age(Mean±SD) | 58.5(±16.4) | 66.2(±13.6) | * |
| Sex (Male) | 43.60 | 61.58 | * |
| ASA Class 1 (%) | 5.97 | 0.26 | * |
| 2 | 42.56 | 6.78 | * |
| 3 | 44.98 | 45.42 | * |
| 4 | 6.32 | 41.12 | * |
| 5 | 0.17 | 6.41 | * |
| Diabetes (%) | 17.19 | 34.23 | * |
| Disseminated Cancer (%) | 2.71 | 5.95 | * |
| Smoker within one year(%) | 18.04 | 24.13 | * |
| COPD(%) | 5.05 | 14.97 | * |
| Ascites(%) | 0.36 | 3.86 | * |
| Congestive Heart<br>Failure(%) | 0.93 | 8.83 | * |
| Hypertension on medication(%) | 49.41 | 73.83 | * |
| Steroid(%) | 4.01 | 9.77 | * |
| Weight Loss(%) | 1.39 | 4.59 | * |
| Bleeding disorder(%) | 4.60 | 17.85 | * |
| Emergency surgery(%) | 10.39 | 37.78 | * |
| Serum Sodium(mEq/L) | 139.1 | 138.0 | * |
| Serum creatinine(mg/dL) | 0.92 | 1.96 | * |
| White blood cell ($\times 10^9/L$) | 8.2 | 11.4 | * |
| Hematocrit(%) | 39.8 | 35.3 | * |
| Platelet($\times 10^9/L$) | 249.2 | 233.1 | * |

\* p<0.001

### Mortality, readmission, length of stay associated with acute kidney injury

**Figure 1** shows a high unadjusted 30-day postoperative mortality rate (37.5%) from 2015-2017 for patients who developed AKI-D compared to those who did not. The rate of 30-day hospital readmission was also higher in these patients (19.7% vs 5.0%). The patients who developed AKI-D also had increased length of hospital stay (18.7 days vs 2.9 days).

**Figure 1.**
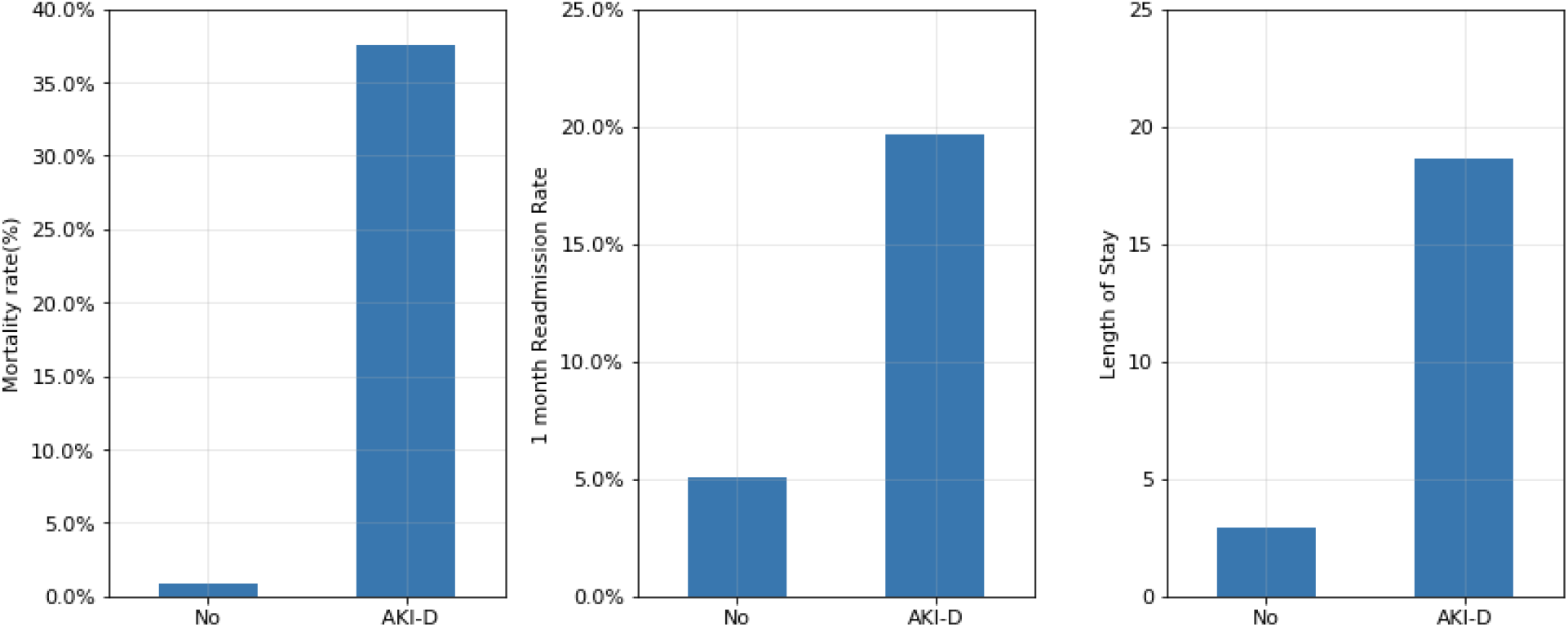
30-day postoperative unadjusted mortality, 30-day hospital readmission rate and length of hospital stay associated with acute kidney injury requiring dialysis.

**Figure 2** illustrates unadjusted 30-day mortality rates according to surgery types from 2015-2017. These varied widely, ranging from 16.2%-53.3%. Although patients undergoing ENT surgery had a high mortality rate, the incidence of AKI-D was low at 0.03%. 1.86% of cardiac surgery patients developed AKI-D with a mortality rate of 37.9%, lower than the rate of 63.7% reported in a previous study from the years 1987-1994. ^2^

**Figure 2.**
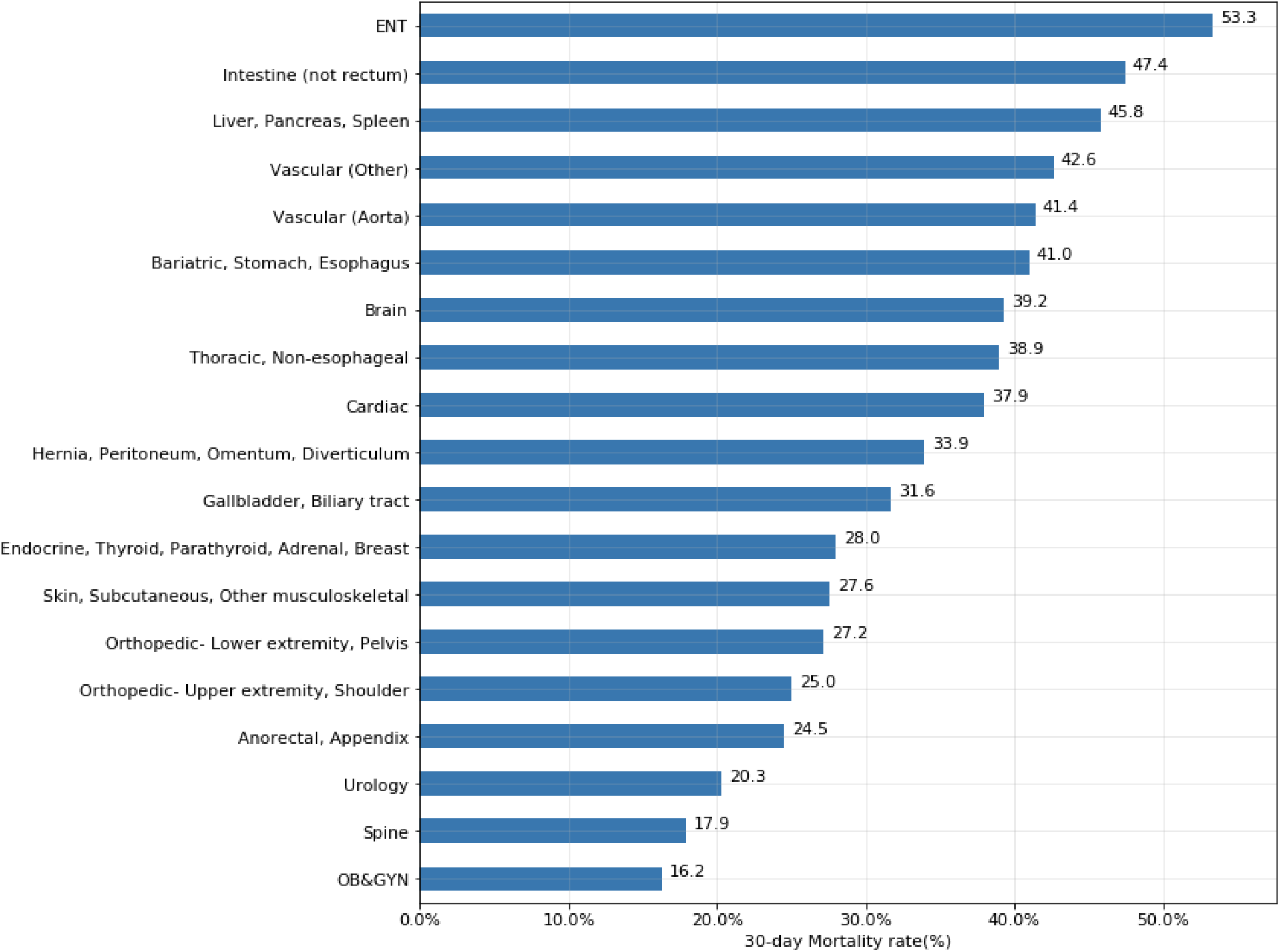
30-day postoperative unadjusted mortality rates of patients who developed AKI requiring dialysis according to surgery types

**eFigure 2** in the **Supplement** shows unadjusted rates of postoperative AKI requiring dialysis according to surgery types. Aorta surgery has the highest risk of AKI-D at 6.05% with a mortality rate of 41.4%. Other surgery types that have higher renal risk include liver, pancreas, spleen surgery (0.82%), intestinal surgery (0.8%), vascular surgery (0.59%) and thoracic surgery (0.48%). Surgery types in lower risk categories include orthopedic upper extremity surgery (0.05%) and spine surgery (0.09%).

### Effect of preoperative renal function, serum sodium and hematocrit

As shown in **Figure 3**, preoperative renal dysfunction was associated with increased unadjusted risk of AKI-D. The risk of acute kidney injury was higher with higher preoperative serum creatinine levels. Anemia also increased the risk of AKI-D, with higher unadjusted rates among patients with hematocrit less than 30% (hematocrit ≤24%: 2.3%, 24.1-30%: 1.4%, >30%: 0.2%). Patients with hyponatremia and hypernatremia had higher incidence of AKI-D (≤130 mEq/L: 1.5%, 130.1-135 meq/L: 0.6%, 135.1-145 mEq/L: 0.2%, >145 mEq/L: 1.0%)

**Figure 3.**
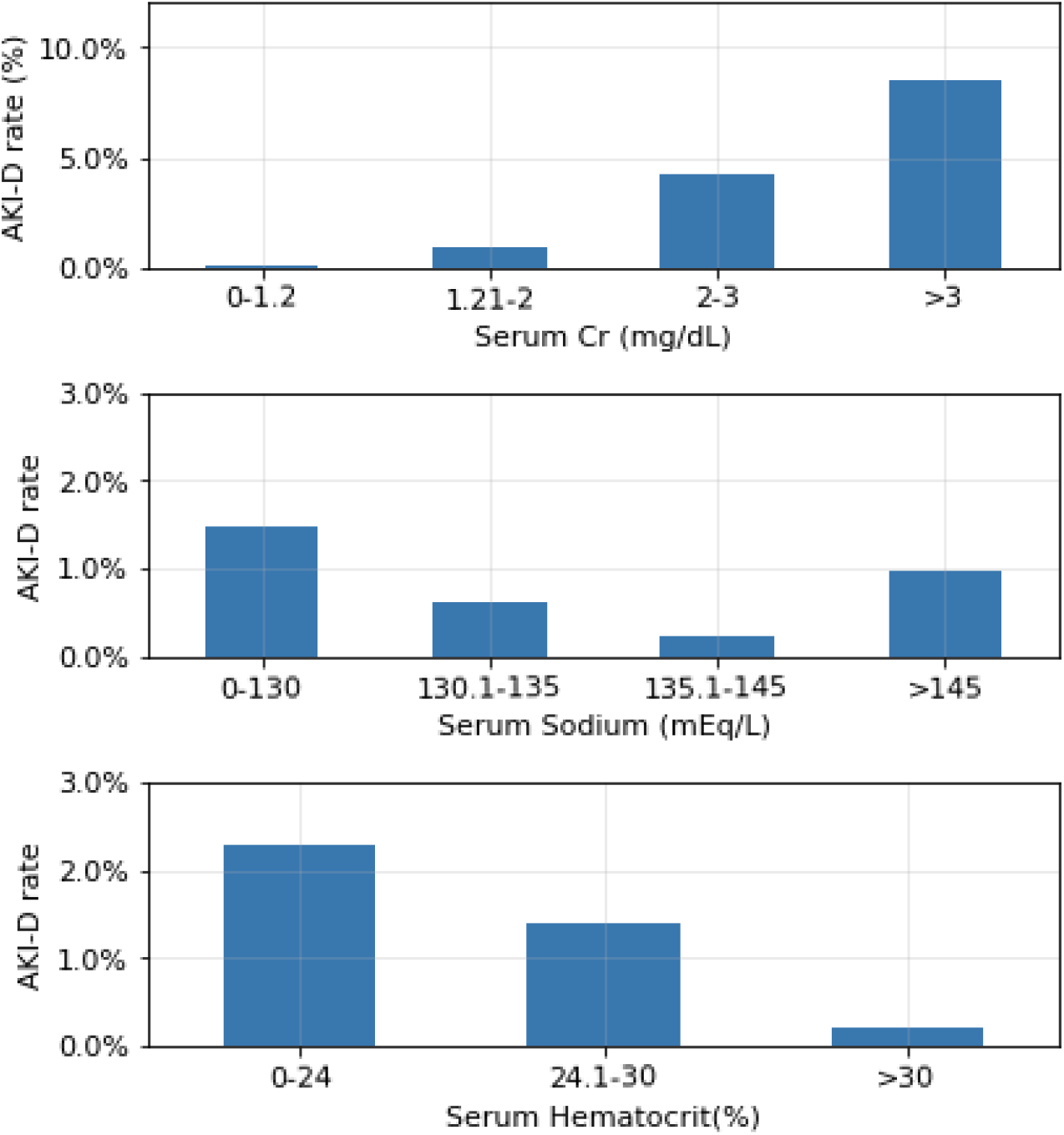
Unadjusted risk of postoperative AKI requiring dialysis according to preoperative serum creatinine, sodium, and hematocrit

### Prediction of acute kidney injury requiring dialysis

**Table 2** lists clinical factors which were strongly associated with the occurrence of postoperative 30-day AKI-D, and were ultimately incorporated into the prediction model. History of congestive heart failure was a significant predictor with an increased adjusted odds of the outcome, OR 2.24 [CI 1.99-2.53, p<0.001]. Diabetes was also associated with an increased risk, OR 2.09 [CI 1.92-2.27, p<0.001] if insulin dependent, OR 1.35 [CI 1.24-1.48, p<0.001] if non-insulin dependent, a finding consistent with a previous study. ^19^ Preoperative condition such as SIRS(systemic inflammatory response syndrome), OR 2.49[CI 2.23-2.78, p<0.001], sepsis, OR 3.26 [CI 2.92-3.64, p<0.001], and septic shock, OR 6.86 [CI 6.04-7.78, p<0.001] were strongly associated with postoperative AKI-D. Increasing ASA class was also significantly associated with AKI-D. Laboratory findings associated with increased risk included preoperative serum sodium and hematocrit. Serum sodium abnormalities in either direction had an increased adjusted risk of the outcome: for serum sodium <=130mEq/L, OR of AKI-D was 1.35 (CI: 1.17-1.55, p<0.001) and for serum sodium 130.1-135mEq/L, OR was 1.21 (CI: 1.12-1.31, p<0.001); for serum sodium >145 mEq/L, OR was 1.47 (CI: 1.23-1.75, p<0.001). Surgery type was also a predictive factor, with the highest rates of AKI-D in patients undergoing aortic surgery, followed by cardiac surgery and surgery of the liver, pancreas, or spleen. Intestinal and other vascular procedures were also strongly associated.

**Table 2.** Predictors of postoperative renal failure requiring dialysis.

|  | Coefficient | SE | P value | Adjusted odds ratio |
| --- | --- | --- | --- | --- |
| Intercept | -9.2277 | 0.122 |  |  |
| Age | 0.0171 | 0.001 | <0.001 | 1.02 (1.015-1.02) |
| Ascites | 0.9321 | 0.09 | <0.001 | 2.54 (2.13-3.03) |
| Congestive heart failure | 0.8077 | 0.061 | <0.001 | 2.24 (1.99-2.53) |
| Emergency surgery | 0.7446 | 0.042 | <0.001 | 2.11 (1.94-2.29) |
| Preoperative acute kidney injury | 1.1624 | 0.064 | <0.001 | 3.2 (2.82-3.63) |
| Diabetes |  |  |  |  |
| Insulin dependent | 0.7346 | 0.042 | <0.001 | 2.09 (1.92-2.27) |
| Non-insulin dependent | 0.3003 | 0.045 | <0.001 | 1.35 (1.24-1.48) |
| Serum Sodium<br>(reference="135.1-145")(mEq/L) |  |  |  |  |
| 0-130 | 0.2974 | 0.073 | <0.001 | 1.35 (1.17-1.55) |
| 130.1-135 | 0.1913 | 0.041 | <0.001 | 1.21 (1.12-1.31) |
| >145 | 0.3826 | 0.091 | <0.001 | 1.47 (1.23-1.75) |
| Serum Hematocrit (%) |  |  |  |  |
| 0-24 | 0.7588 | 0.074 | <0.001 | 2.14 (1.85-2.47) |
| 24.1-30 | 0.6566 | 0.042 | <0.001 | 1.93(1.78-2.09) |
| Preoperative Sepsis<br>(reference = No) |  |  |  |  |
| SIRS | 0.9105 | 0.056 | <0.001 | 2.49 (2.23-2.78) |
| Sepsis | 1.182 | 0.057 | <0.001 | 3.26 (2.92-3.64) |
| Septic Shock | 1.925 | 0.064 | <0.001 | 6.86 (6.04-7.78) |
| Surgery type (reference =<br>anorectal, appendix) |  |  |  |  |
| ENT | 0.2652 | 0.371 | 0.475 | 1.30 (0.63-2.7) |
| Bariatric, stomach, esophagus | 1.4485 | 0.127 | <0.001 | 4.26 (3.32-5.46) |
| brain | 0.8844 | 0.21 | <0.001 | 2.42 (1.61-3.65) |
| cardiac | 2.9198 | 0.141 | <0.001 | 18.54(14.06-24.44) |
| Endocrine, thyroid,<br>parathyroid, adrenal, breast | -0.4172 | 0.203 | 0.040 | 0.66 (0.44-0.98) |
| Gallbladder, biliary tract | 0.5608 | 0.14 | <0.001 | 1.75 (1.33-2.31) |
| Hernia, peritoneum, omentum,<br>diverticulum | 0.934 | 0.121 | <0.001 | 2.55 (2.01-3.23) |
| Intestine (not rectum) | 1.6948 | 0.111 | <0.001 | 5.45 (4.38-6.77) |
| Liver, pancreas, spleen | 2.577 | 0.126 | <0.001 | 13.16 (10.28-16.84) |
| OBGYN | 0.1946 | 0.173 | 0.262 | 1.22(0.87-1.71) |
| Orthopedic lower extremity,<br>pelvis | 0.7366 | 0.118 | <0.001 | 2.09 (1.66-2.63) |
| Orthopedic upper extremity,<br>shoulder | 0.0239 | 0.261 | 0.927 | 1.02 (0.61-1.71) |
| Skin, subcutaneous, other<br>musculoskeletal | 0.9193 | 0.13 | <0.001 | 2.51 (1.95-3.23) |
| Spine | 0.6338 | 0.148 | <0.001 | 1.89 (1.41-2.52) |
| Thoracic (non-esophageal) | 1.9322 | 0.149 | <0.001 | 6.91 (5.16-9.25) |
| Urology | 1.5805 | 0.124 | <0.001 | 4.86 (3.81-6.19) |
| Aorta | 4.0768 | 0.128 | <0.001 | 58.96 (45.91-75.72) |
| Vascular | 1.8613 | 0.117 | <0.001 | 6.43 (5.11-8.1) |
| Serum Creatinine(mg/dL) | 0.4055 | 0.009 | <0.001 | 1.5 (1.47-1.53) |

These predictors were used to develop a model for AKI-D as described in the methods section. Brier score for the training cohort was 0.003. The model has a training AUC of 0.89. **eFigure 3** in the **Supplement** shows the calibration plot of the prediction model which is well matched with a 45-degree line.

### Validation of the renal risk model

The trained model was tested by applying the model to a validation cohort of 811,778 from 2017. The prediction model has excellent predictive power with AUC of 0.90. 5-fold cross validation from the training cohort also showed high average AUC (0.89). **Figure 4** shows AUC of model validation.

**Figure 4.**
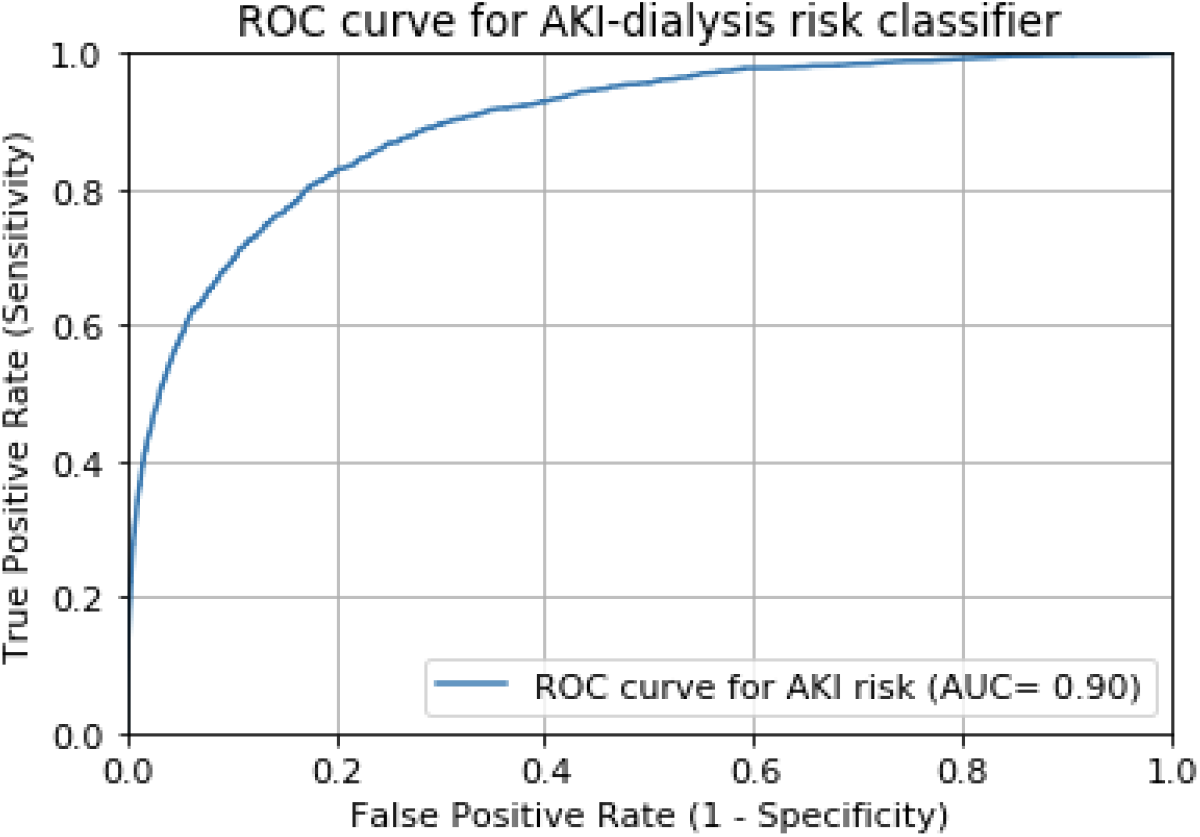
Predictive model performance with validation cohort for postoperative AKI requiring dialysis

### Examples of Renal Risk Calculator

A calculator for AKI-D risk was programmed from coefficients and intercept from multivariate logistic regression and Scikit-learn machine learning library. ^20^. The following examples demonstrate the bedside applicability of this model as well as the impact of various clinical factors on the risk of the outcome. The mobile web model is available at https://renalrisk.herokuapp.com/. The mobile-web model is shown in **eFigure 4** in the **Supplement**.

#### Hypothetical case 1

67 year old male with a history of congestive heart failure and insulin dependent diabetes mellitus undergoing emergent hip replacement surgery. No ascites. Patient had preoperative acute kidney injury and SIRS (systemic inflammatory response syndrome). Preoperative laboratory values: serum creatinine 2.5 mg/dL, hematocrit 24%, sodium 128 mEq/L. **Probability of AKI requiring dialysis (AKI-D) within 30 days after surgery: 28**.**36%**

#### Hypothetical case 2

67 year old male with a history of congestive heart failure and insulin dependent diabetes mellitus undergoing liver (hepatectomy) surgery. Patient had no preoperative acute kidney injury and SIRS. Preoperative laboratory values: serum creatinine 2.5 mg/dL, hematocrit 32%, sodium 133 mEq/L. **Probability of AKI-D within 30 days after surgery: 5**.**9%**

## Discussion

We have developed a risk model for postoperative AKI-D in patients undergoing broad types of surgery. This model uses simple, easily available clinical data to deliver excellent predictive ability for the outcome, with a validation AUC of 0.90.

Our study has strengths compared to prior models to predict postoperative renal failure. A prior machine learning algorithm to predict postoperative renal failure has been described with lower predictive ability (AUC ranging from 0.797 and 0.858) and requiring a complex set of EHR-extracted data.^21^ There are also previously described machine learning prediction models for acute renal failure in hospitalized patients, not specific to the postoperative population and often relying on complex data extracted from the EHR.^22 23 24 25^

Other strengths of this study include the very large (2,299,502 patients) and recent (2015-2017) cohort used to build our model. The study population includes various surgery types including cardiac surgery, and the risk model was validated across surgery types.

Another strength of this model is the inclusion of potentially modifiable variables such as preoperative serum sodium and hematocrit, which are potential targets for preoperative optimization. Identifying modifiable pre-op risk factors for renal failure is an unmet clinical need. Whether the changes of these modifiable factors improve the renal outcome will require further investigation.

A limitation of our model is an inability to account for multiple factors as is possible in an EHR-integrated model, for example zip code as a predictor of socioeconomic factors. The simplicity of our model was an intentional choice, making it useful in a variety of settings without requiring institutional investment in EHR integration. Additionally, the large dataset used to develop and validate the model portends a high degree of external validity. This could be further tested by using this model at institutions that do not participate in NSQIP and evaluating its performance in that population, which will assure the generalizability of the model.

Another limitation pertains to the clinical use of this model. Though certain factors investigated are modifiable (for example, preoperative hematocrit and sodium), this analysis cannot determine causality, or the impact of modifying these factors on ultimate risk of AKI-D. An individual with a sodium <130 mEq/L can be counseled on an elevated postoperative risk of AKI-D, but it is not known if correcting the sodium prior to surgery will be protective.

This analysis shows that preoperative renal function, older age, congestive heart failure, diabetes and anemia are all significantly associated with the development of postoperative AKI-D. A clinical model incorporating 11 readily-available patient factors can be easily used by the clinician at bedside and has high predictive ability for this outcome, making it a useful and individualized tool for shared surgical decision-making. Further study will be needed to see if optimizing modifiable predictors can improve renal outcomes.

## Data Availability

Data is available at American College of Surgeons (ACS-NSQIP).

## Acknowledgments

*The American College of Surgeons National Surgical Quality Improvement Program and the hospitals participating in the ACS NSQIP are the source of the data used herein; they have not verified and are not responsible for the statistical validity of the data analysis or the conclusions derived by the authors*.

